# Machine learning and MRI-based diagnostic models for ADHD: are we there yet?

**DOI:** 10.1101/2020.10.20.20216390

**Authors:** Yanli Zhang-James, Ali Shervin Razavi, Martine Hoogman, Barbara Franke, Stephen V Faraone

## Abstract

Machine learning (ML) has been applied to develop magnetic resonance imaging (MRI)-based diagnostic classifiers for attention-deficit/hyperactivity disorder (ADHD). This systematic review examines this literature to clarify its clinical significance and to assess the implications of the various analytic methods applied. We found that, although most of studies reported the classification accuracies, they varied in choice of MRI modalities, ML models, cross-validation and testing methods, and sample sizes. We found that the accuracies of cross-validation methods inflated the performance estimation compared with those of a held-out test, compromising the model generalizability. Test accuracies have increased with publication year but were not associated with training sample sizes. Improved test accuracy over time was likely due to the use of better ML methods along with strategies to deal with data imbalances. Ultimately, large multi-modal imaging datasets, and potentially the combination with other types of data, like cognitive data and/or genetics, will be essential to achieve the goal of developing clinically useful imaging classification tools for ADHD in the future.

## Introduction

Clinicians diagnose ADHD by evaluating symptoms of hyperactivity, impulsivity, inattention, and impaired functioning across settings. The diagnosis of ADHD shows considerable levels of concurrent and predictive validity in its clinical features, course, neurobiology, and treatment response [1, 2]. Nevertheless, concerns about diagnostic accuracy persist. Some suggest that the current method of diagnosing ADHD is too subjective and leads to over-diagnosing ADHD in the community [3, 4]. Psychiatric diagnoses have been called “subjective” because they rely on clinician evaluation of responses from patients, parents, and/or informants. Other studies have raised concerns about the under-diagnosis of ADHD [5, 6], especially in girls and women, which suggests biases in applying the current diagnostic algorithm. Another issue is the misdiagnosis of ADHD as being another disorder. When this occurs, patients may be exposed to unnecessary treatments and will continue to struggle with the many impairments associated with ADHD. Those who have ADHD and are not diagnosed with the disorder will continue to have impaired functioning leading to increased risks for other health and social problems [7-11].

In response to these concerns, researchers have sought to develop objective measures to diagnose ADHD or to monitor the course of ADHD symptoms during treatment. Much research has examined peripheral biochemical markers in differentiating ADHD and control patients, such as (Norepinephrine (NE), 3-Methoxy-4-hydroxyphenylethylene glycol (MHPG), monoamine oxidase (MAO), zinc, and cortisol [12, 13]. NE, MHPG, MAO, b-phenylethylamine, and cortisol are also somewhat predictive of response to ADHD medications. Meta-analysis also shows that peripheral measures of oxidative stress differ between ADHD and control participants [14]. Electroencephalographic (EEG) [15], actigraphic [16], and eye vergence measurements [17], as well as interactive gaming behaviour [18] were also examined as ADHD biomarkers. Neuropsychological tests [19], particularly continuous performance tests (CPTs) [e.g. 20, 21, 22] have been evaluated in many studies. In recent years, genetic markers in the form of polygenic risk scores also have shown some predictive ability of diagnosis and prognosis of ADHD [23-25].Many of these prior studies show group differences but do not present diagnostic accuracy statistics. A clinically useful biomarker should have at least 80% sensitivity and 80% specificity. They should also be reliable, reproducible, inexpensive, non-invasive, easy to use, and confirmed by at least two independent studies. These criteria were defined by work of the task force on biological markers by the World Federation of ADHD [26]. None of the measures examined by them met these criteria for clinical utility [26].

Prior structural MRI (sMRI) studies have consistently reported alterations in frontal, parietotemporal, cingulate, cerebellum, basal ganglia, and corpus callosum regions [27-33]. Studies of the largest ADHD sMRI dataset from the Enhancing Neuro Imaging Genetics Through Meta-Analysis (ENIGMA) consortium’ s ADHD Working Group reported the significant volumetric reductions in intracranial volume, amygdala, caudate nucleus, nucleus accumbens, hippocampus, and cortical surface areas from many regions in children with ADHD [34, 35]. These regions have also been implicated in functional MRI (fMRI) studies showing altered brain connectivity and activation in the fronto-striatal, fronto-parietal and fronto-temporal-parietal circuits, as well dorsal anterior cingulate cortex in ADHD brains [36-38]. Studies have also examined the developmental trajectories of these anatomical and functional alterations across the lifespan finding initial delays that are followed by apparent normalization [27, 29, 31].

These findings encouraged efforts to develop objective diagnostic tools for ADHD using MRI data. Early studies used standard statistical methods such as discriminant analysis with very small sample sizes [39-41]. For example, a discriminant analysis reported by Semrud-Clikeman et al. [41] included 10 participants in each of three diagnostic groups: developmental dyslexia, ADHD or control. Zhu and colleagues’ discriminant analysis classifier assessed 9 ADHD and 11 typically developing boys. Although high predictive accuracies were reported in these studies (85∼ 87%), it is difficult to evaluate how well those models would generalize given the small samples and lack of replication samples.

The ADHD-200 Global Competition [42] challenged researchers to develop an MRI-based diagnostic classifier for ADHD. It provided a dataset of 776 children, adolescents and young adults (7-21 years old, 63% healthy controls, 37% ADHD) from eight sites. Fifty teams from around the world joined the competition with 21 final submissions. Machine learning models predominated. Due to the large sample size, the consortium was able to set aside a test set that was not used for model selection and development. The competition results were judged by the performance on the test set only. This contrasts with previous studies with small sample sizes, where a held-out test set was not available. The ADHD-200 winning team used an ensemble model which achieved a 61% accuracy with 21% sensitivity and 94% specificity using both structural and resting state-fMRI data along with the demographic predictors [43]. The accuracy, although considerably lower than previously reported high accuracies, was one of the first in an independent, held-out test set. Despite the modest accuracy, the ADHD-200 competition re-kindled enthusiasm for developing imaging-based diagnostic classifiers for ADHD. The publicly available dataset has become the main data source driving the machine learning model development for ADHD. Since the competition in 2012, we have seen a steady increase in the number of publications applying machine learning classifiers to ADHD. Thirty-one additional published studies have used either the whole or part of the ADHD-200 dataset (Supplementary Figure 1 and Table 1).

**Table 1.**
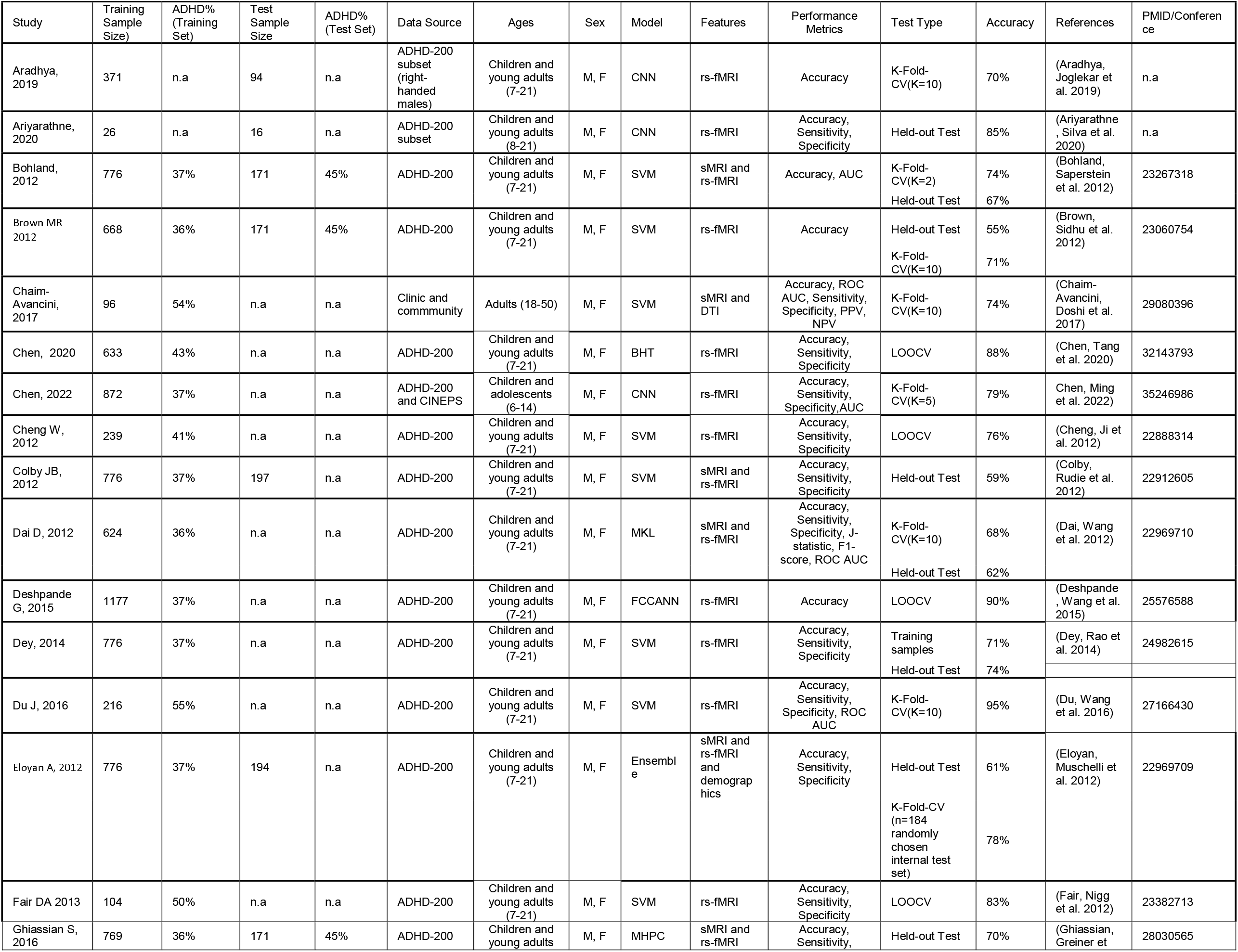
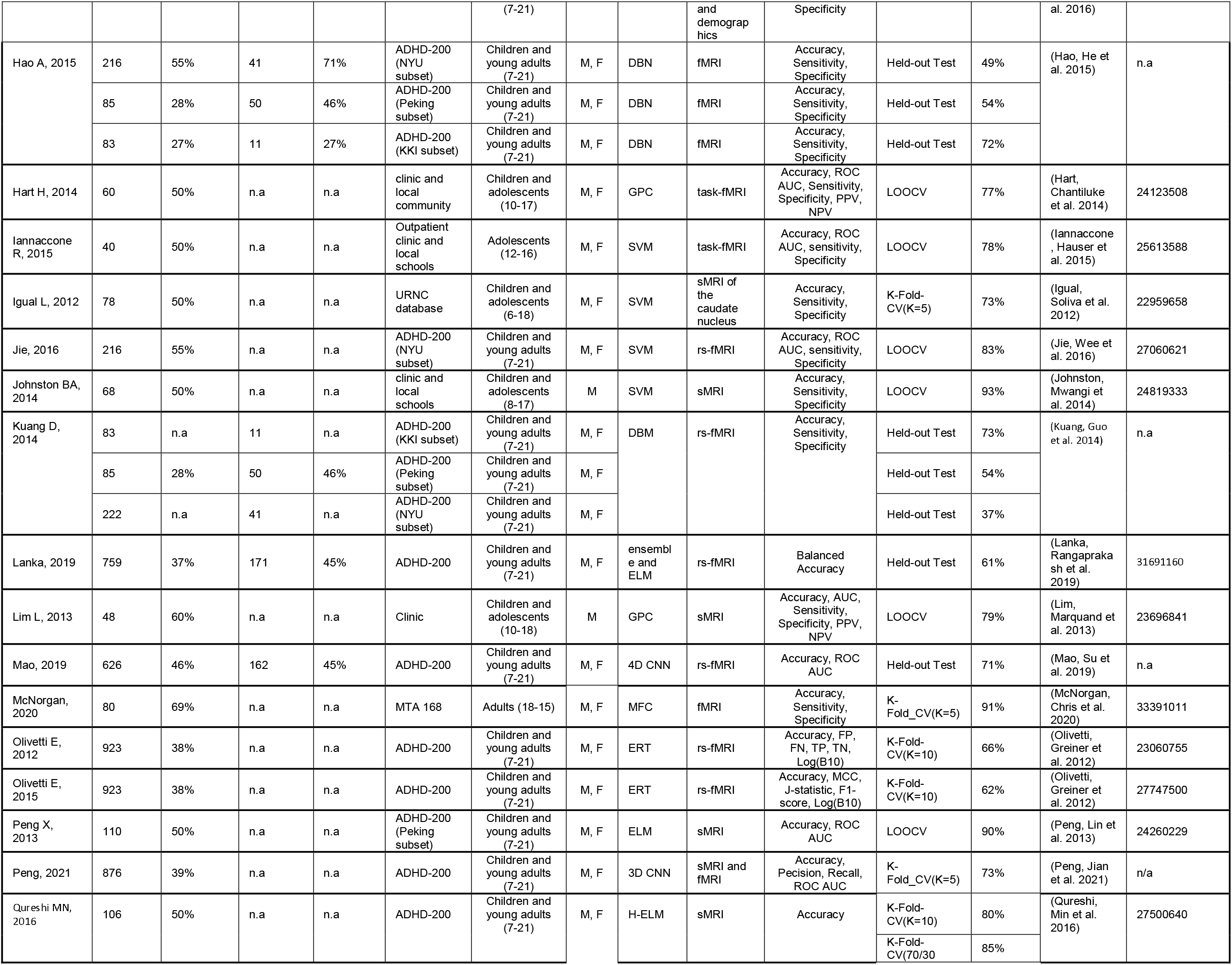
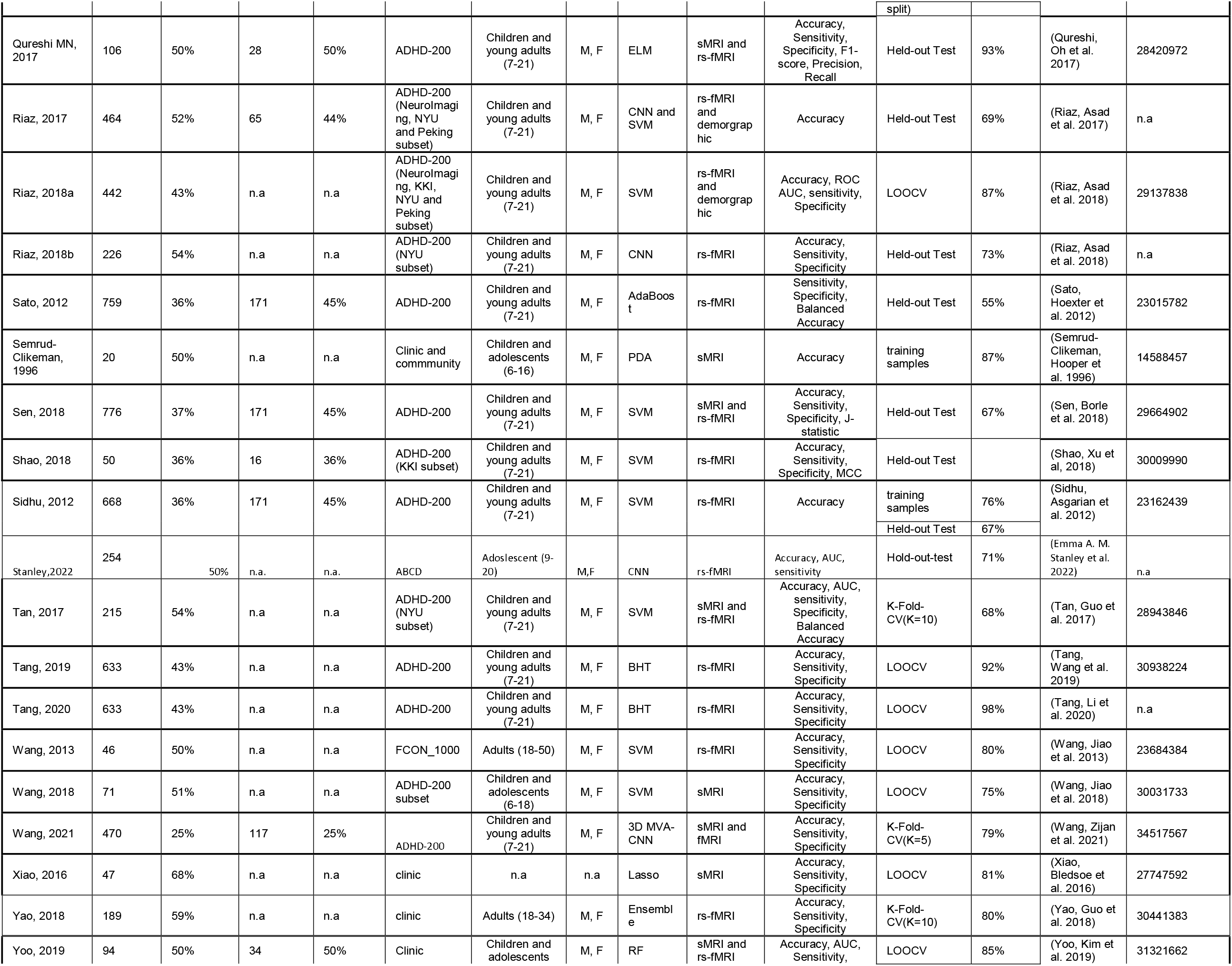
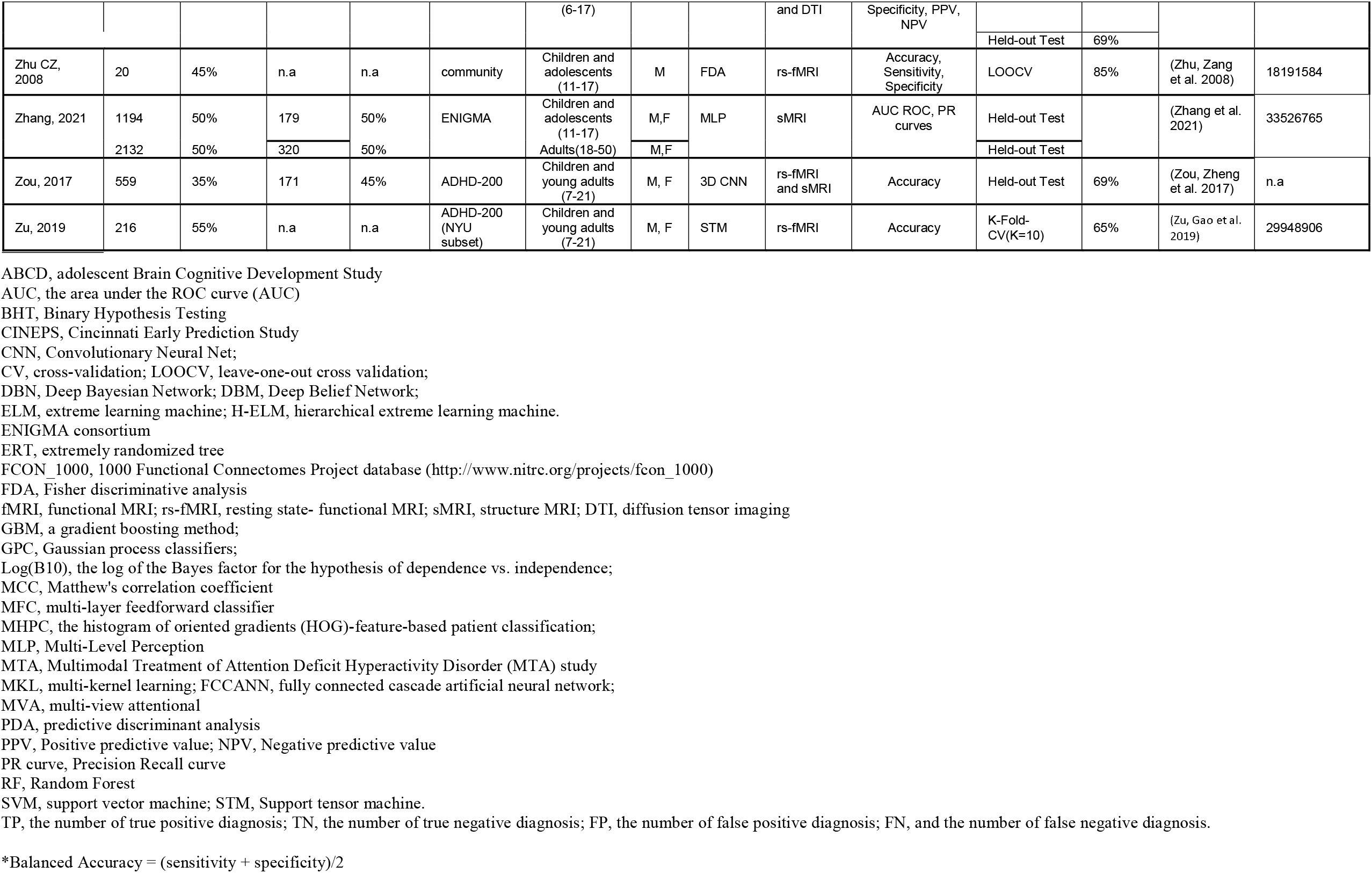
Machine Learning Literature on ADHD Neuroimaging Data.

This systematic review examines the prior literature applying ML to MRI data in ADHD to clarify the clinical significance of findings and to assess the implications of the various analytic methods applied. We discuss the progress made over the years as well as lessons and methodological issues that we learned from this body of work. We hope to provide a roadmap for future studies that aim to overcome these issues and achieve clinically useful models for diagnosing ADHD.

## Methods

A literature search on MRI-based diagnostic classifiers for ADHD using key words (“ADHD” AND “MRI” AND (“Machine learning” OR “Classi*”)) and examining their references identified 55 studies in total (up to July 1st, 2022, Pubmed, Embase and Google).

Supplementary Figure 2 shows the article selection procedure in a PRISMA diagram. The eligible studies applied statistical or machine learning classifiers using MRI data to differentiate participants with ADHD from controls. Table 1 lists the selected studies along with the performance of their best models. If a study dealt with multi-class classification, for example, having ADHD, ASD and control groups, only the two-class classification accuracies involving ADHD vs the control groups were examined in this review. We used percent correct (accuracy) to compare results across studies because it was available for most of the papers. Studies that met the classifier criteria but did not report an accuracy statistic or other metrics that can be used to compute accuracies were not included in our quantitative analysis. If a study reported multiple models, only the model which had the highest accuracies was included in Table 1.

We extracted and examined study characteristics, including machine learning model types, MRI data modality, cross-validation and testing methods, training sample size, training set class ratio (the ratios of ADHD vs Control participants’ numbers), data source, dataset age and sex compositions and publication years, etc. We grouped machine learning models to three categories: support vector machine (SVM), convolutional neural networks (CNNs), and others. We assigned studies with a training set class ratio between 0.4 ∼0.6 as “balanced” (i.e., nearly equal), and those with higher or lower ratios as “unbalanced”. Nine studies used various methods to balance demographic differences between the ADHD and control groups. These were assigned as “balanced”, even if their original class ratio was outside of the balanced range [44-51] We reported the age and sex groups, as well as the minimum and maximum age range of the dataset. For the ADHD-200 samples, the overall age range was used if a specific subset was used but age information was not provided. Minimum and maximum values of age were derived for studies that reported mean and standard deviation of the ages.

We also classified studies based on the methods they used to evaluate model performance and generalizability. Two methods were used. The held-out test set method evaluates model performance on data that were set aside, i.e., they were not used during model estimation and training. Because this method requires a large sample, many studies resort to cross-validation (CV) method to assess model performance. CV methods randomly re-sample examples to be set aside during model fitting. The most commonly used versions are the leave-one-out CV (LOOCV) [46, 47, 52-54] and K-Fold-CV (where K is often = 10, 5, or 2) [44, 55-57]. For example, in 10-fold CV, the original dataset is partitioned into 10 equal sub-samples or “folds”. For each iteration of model estimation nine of the subsamples are used to estimate model parameters and the left-out fold is used to estimate model accuracy. The left-out fold changes from iteration to iteration. For LOOCV, one sample is left out for testing while all the others are used for training or model fitting. In either situation, the process is repeated until all samples have been used in both the training and test sets. The CV accuracy is estimated by averaging over all iterations of CV accuracies. Although CV samples were not used during the model training/fitting at each iteration, they are, nevertheless, used as training examples in other iterations.

Our main objective was to understand how study features influenced model accuracy. We used likelihood-ratio (LR) test assisted variable selection in combination with multivariate linear regression to quantitatively evaluate if these features predicted model accuracy. The variable selection algorithm implemented in STATA16’ s *gvselect* command computes both the Akaike’ s [58] information criterion (AIC) and Schwarz’ s [59] Bayesian information criterion (BIC) [60]. We performed the variable selection and linear regression modeling for all the studies combined, as well as separately for the K-Fold-CV, LOOCV, and held-out test groups. Training sample size was primarily examined as a continuous variable. However, we also classified sample sizes as small (<300) or large (>300) to compare the variability of their accuracy estimates using Levene’ s robust test statistic [61]. In addition to the quantitative analysis, we also qualitatively reviewed the relevant study characteristics if a quantitative analysis was not possible.

## Results

Among all the studies included, over half the studies (N=31, 56%) reported only CV results without a held-out test set. Forty-two percent (N=23) used a Hold-out test sample to evaluate classifier performance. All but one of the 23 studies used the ADHD-200 samples. Among the studies that reported held-out test results, six also reported CV results. Figure 1 shows that the 16 studies using K-Fold-CV and the 17 studies using LOOCV reported, on average, higher accuracies than studies using held-out tests (F_(2,55)_ = 34.52, p < 0.001).

**Figure 1.**
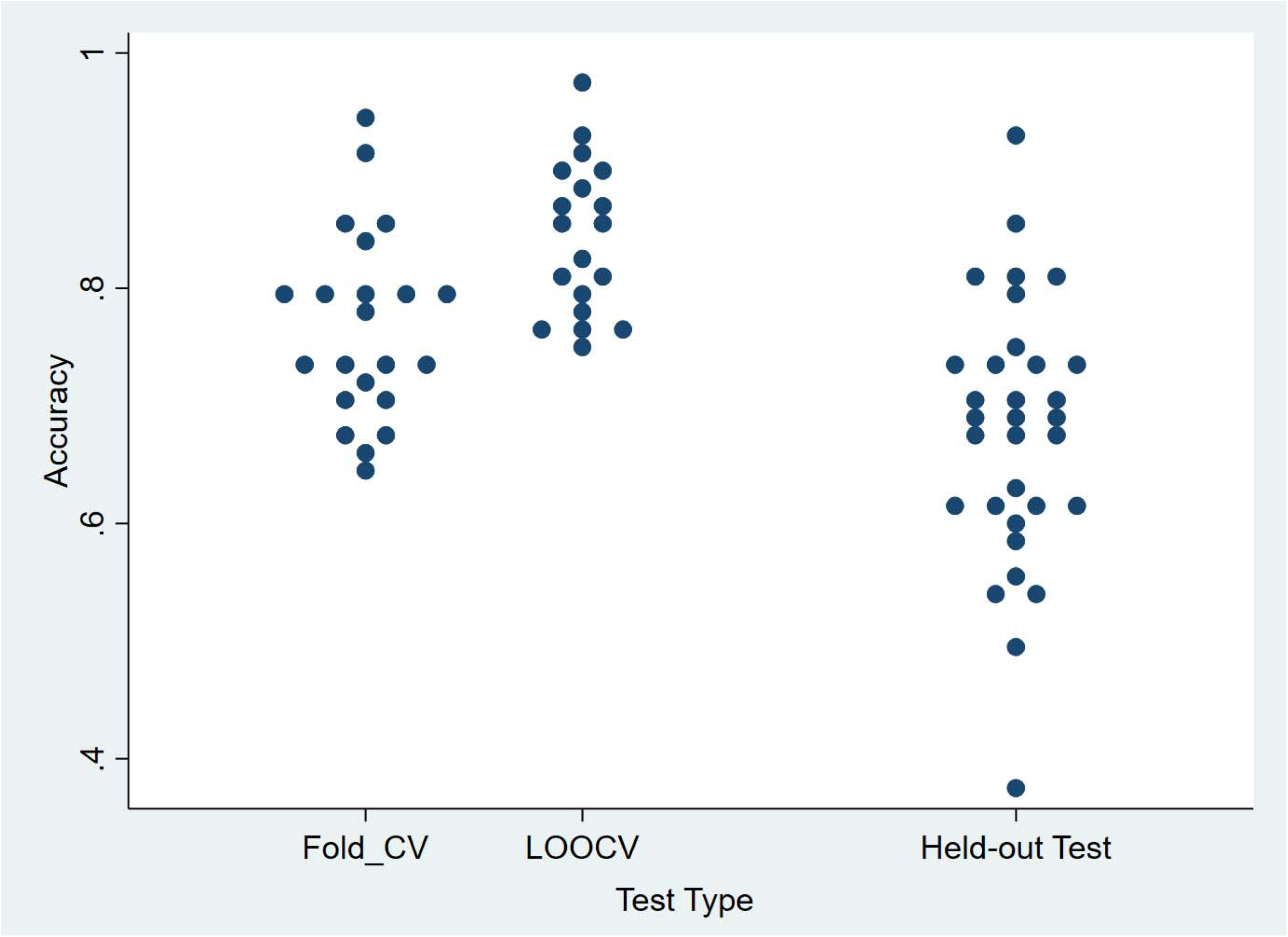
Best prediction accuracies reported in each study for each type of the available tests: K-Fold-CV, LOOCV or held-out tests.

Excluding a 2021 outlier study with the largest sample size and low accuracy[51], the accuracy estimates overall increased in later publication years (F_(1, 54)_=5.15, p =0.027 Figure 2). There was no significant change of reported accuracies over the years in studies using the K-Fold-CV methods (Figure 2 Left). The increase was primarily driven by studies using LOOCV and Hold-out test sets that showed a statistically significant increase over time (F_(1, 17)_=12.63, p=0.0024, F_(1, 22)_=4.61, p=0.043, Figure 2 Middle and Right).

**Figure 2.**
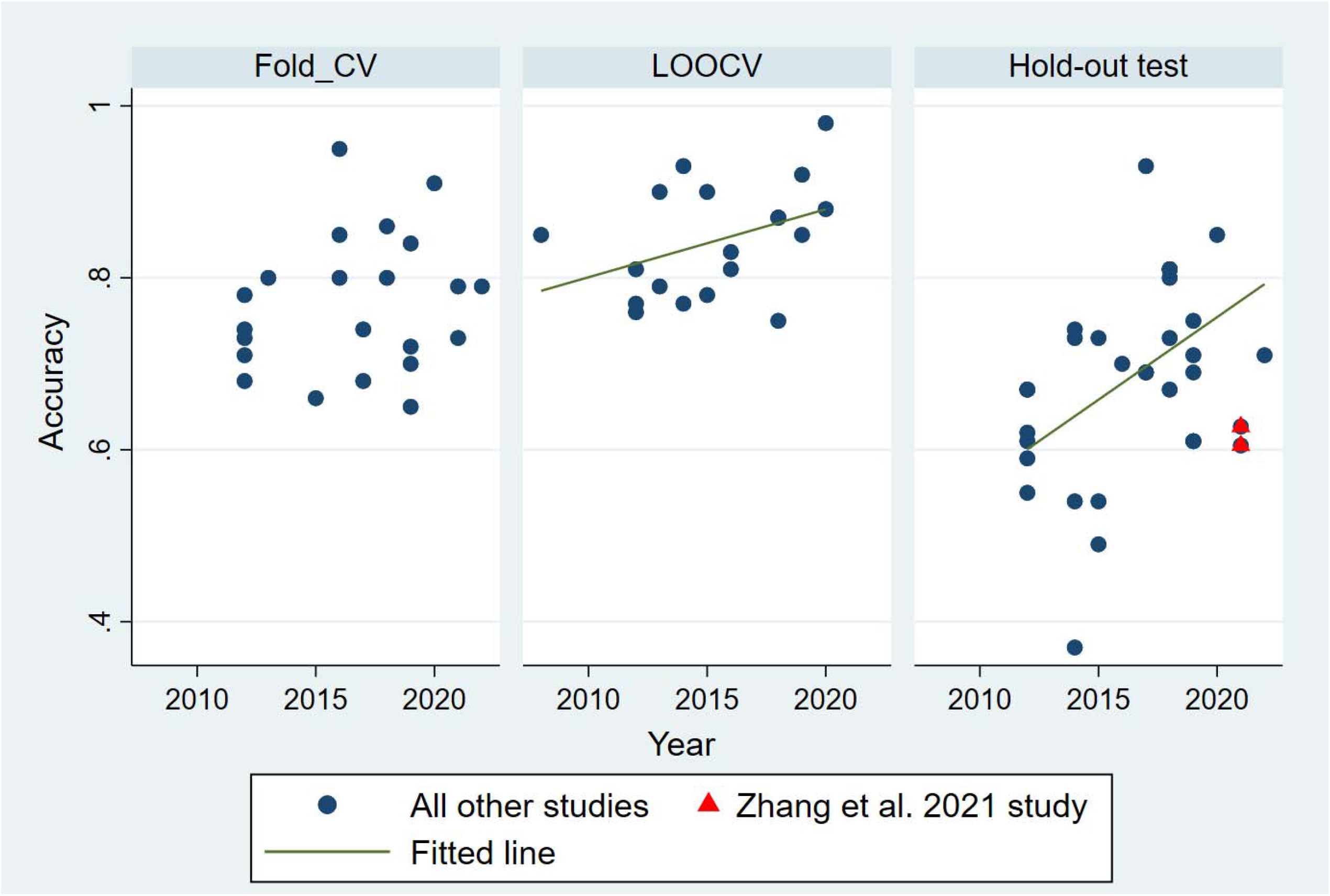
Accuracy in studies published over the years. Zhang et al. 2021 study is highlighted with red triangles to signify its outlier status due to reported the large sample size and low accuracy.

Training sample size, overall, was not significantly associated with accuracy (Figure 3 Left and Right), either as a continuous or categorical variable. However, it negatively predicted accuracies in the LOOCV group (F_(1, 17)_ = 10.15, p = 0.005, Figure 3 Middle). Studies with large samples had lower mean accuracies than studies with small studies (72% vs 77% mean accuracies, t=1.79, p = 0.038). Furthermore, the accuracy results from small studies were more variable than those from large studies in the held-out test group (Levene’ s robust test statistic W0 _(1, 30)_ = 6.58, p = 0.015). The variance differences between large and small samples were not statistically significant for either the K-Fold-CV or LOOCV.

**Figure 3.**
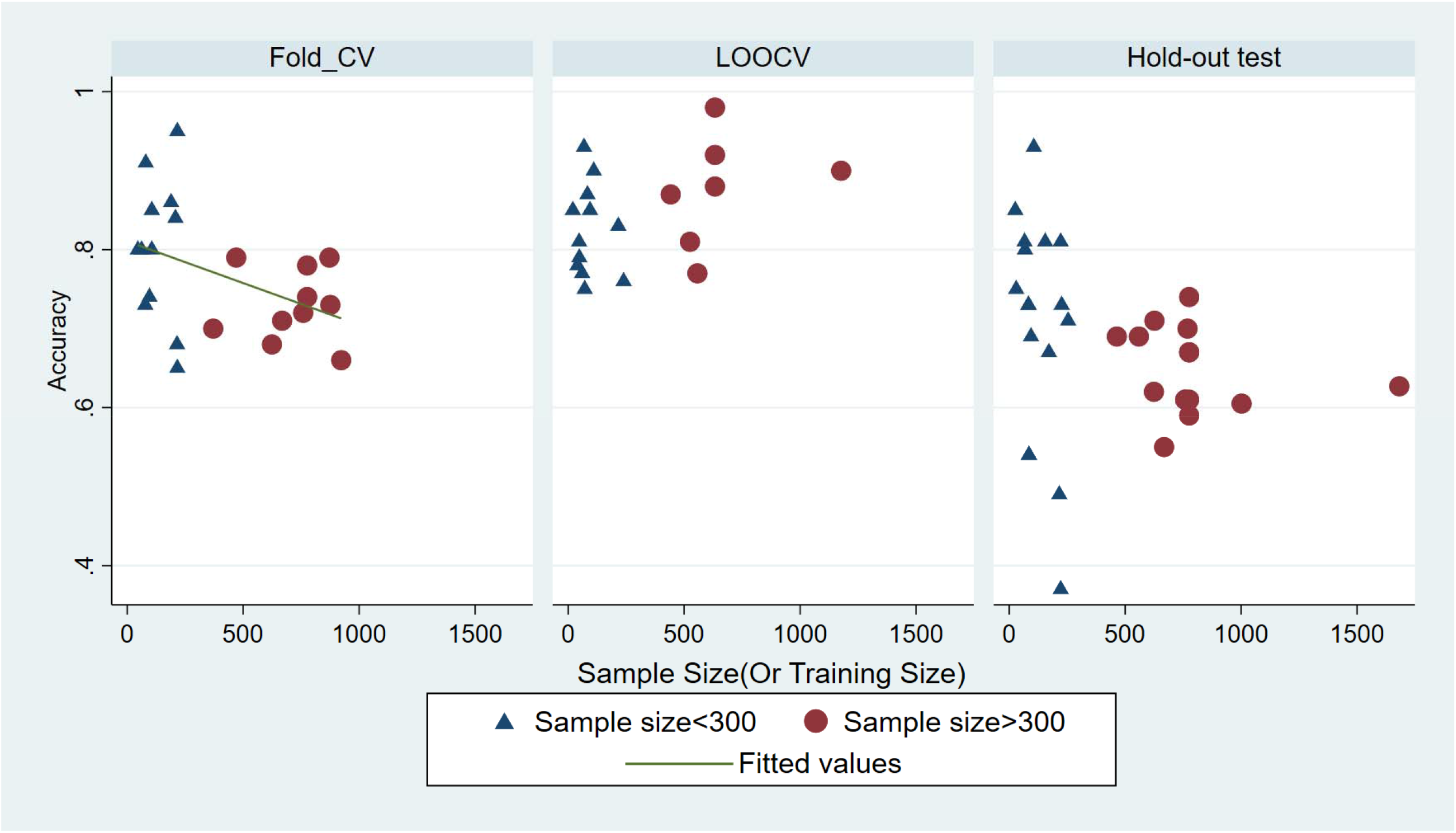
Accuracy vs training sample size. Sample sizes <300 were labeled as triangle and >300 are labeled as circle. The fitted line between accuracy and sample size were plotted for each test type.

Twenty-four studies (49%) used a training dataset that had severely imbalanced classes. Nine of those studies applied data balancing methods to compensate for the class imbalance and are grouped as balanced studies. Class-balanced studies reported higher accuracies for both the K-Fold-CV (F_(1, 19)_ = 6.55, p = 0.02)and LOOCV (F_(1, 17)_ = 36.02 and p < 0.0001). However, the balanced studies in the K-Fold-CV group were mostly small studies with the exception of three studies (Supplementary Figure 3B); we could not differentiate whether the higher accuracy was due to the negative relationship with sample size or the benefit of data balance. The higher accuracies in the balanced LOOCV group were related to sample size as only the large group (>300 samples) showed a significant relationship between accuracy and the balanced criterion (F_(1, 5)_ = 44.81, p = 0.0011).No statistical difference was found for either accuracies or training sample size between the balanced and unbalanced studies in the held-out test group.

Because the ADHD-200 dataset was the main data source, most studies (N=29) used resting-state fMRI data (rs-fMRI), or rs-fMRI in combination with sMRI data (multi-modal, N=16). Only eight studies used sMRI data, and only two used task-based fMRI data. There were no significant differences in sample size across the different feature types (Supplementary Figure 4A). However, except for the two task-based fMRI studies, which both used LOOCV and reported significantly lower accuracies than other MRI modalities (t=-23.3, p<.0001), there was no difference in reported classification accuracies observed among the sMRI, rs-fMRI, or multi-modal studies (Supplementary Figure 4B).

The ADHD-200 dataset has a mix of children, adolescents and young adults (age 7-21). Ten other studies focused only on children and/or adolescents (under age 18). Only five studies examined classification models for older adults [45, 51, 62-64]. Overall, the difference in accuracy across the three types of age compositions was not significant (F_(2,55)_=1.74, p=0.19).

Most studies used a mixture of male and female participants. Four studies only included boys [40, 63, 65, 66]. These four reported significantly higher classification accuracies than all other studies that used a mixture of males and females (F_(1,55)_ =32.14, p < 0.0001). However, all four were small studies (n=20∼189). Three reported LOOCV and one reported 10-Fold-CV accuracies.

Across all studies, the most frequently used model was the support vector machine (SVM). It was used in 20 (36%) studies. SVM, and most other ML models cannot directly analyze images. Instead, they analyze some transformation of images such as regional volumes or cortical thickness. In contrast, convolutional neural networks (CNNs) can analyze images directly and thus have access to all the information available. Only in recent years (2017-2022) have studies applied CNN methods to MRI images (N=11). We did not find any statistically significant differences between the accuracies reported with SVM, CNN, or other models for ADHD (F_(2, 55)_ = 0.01, p = 0.91, Supplementary Figure 5).

## Discussion and Qualitative Review

Our quantitative analysis of prediction accuracy for ADHD revealed several significant findings. First, accuracies based on K-Fold-CV or LOOCV were significantly higher than those reported using held-out tests, which suggests that CV methods may over-estimate model performance. Second, we found greater variability of test accuracies reported in studies with small sample sizes than those of larger sample sizes and an inverse relationship of sample size and K-Fold-CV accuracies. Third, estimates of accuracy increased with publication year. This was driven primarily by the Hold-out and the LOOCV test-groups. Since sample size has been roughly the same since 2012, with the exception of a 2021 study [51], the increasing accuracy over time could be due to several design features: 1) the use of more sophisticated models (such as deep neural networks and CNN models), 2) improved methods of data balancing and data augmentation, and 3) use of feature selection, feature space reduction methods or different MRI data modalities. However, our analysis cannot conclusively clarify if any or all of above attributed to the increase of accuracies. We discuss the implications of these findings and provide further review of some study characteristics that were not examined in our quantitative analysis.

### Cross-Validation vs Held-out Test Set

In the CV approach, the validation samples used to estimate accuracy are not used during the model training/fitting at each iteration. They are, nevertheless, used as training examples in other iterations. Moreover, because there are many iterations, the validation set can influence parameter estimates. In contrast, the held-out test method uses a test set that was never used during model training. As a result, CV accuracies have been shown to overestimate test set accuracy when both are available [56, 57]. Our results confirm the inflation of accuracy by K-Fold-CV or LOOCV. Held-out test accuracy is a better indicator of model performance with unseen samples.

More than half the studies (N=31, 56%) reported only CV results without a held-out test set. An earlier review reported 13% of ADHD neuroimaging (including MRI and electroencephalographic) studies consisted of “circular analysis”, where independent test sets were not used [67]. Our results are more similar to what Kriegeskorte et al. [68] had estimated, 42%∼56% of studies consisted of “circular analysis”, based on all fMRI studies published in five prestigious journals (Nature, Science, Nature Neuroscience, Neuron, Journal of Neuroscience) in 2008. Nevertheless, our review highlights the importance of building a large dataset through collaborations and open data sharing as we pointed out that the majority of the studies that were able to afford a held-out test were those that used the ADHD-200 dataset.

### Sample Size

Machine learning, particularly deep learning, often requires large sample sizes due to the large number of parameters and hyperparameters that a model needs to learn. However, many neuroimaging studies of ADHD used very small sample sizes. In our small sample group, the sample size ranged from 20 to 239 (average sample size 112). Small sample sizes can lead to model overfitting and overestimates of accuracy [69, 70]. In our review, this effect was reflected in the large variability of accuracies in the CV studies. Indeed, some of the highest and lowest test accuracies were reported in studies with extremely small sample sizes. None of the studies reviewed here used a learning curve analysis to assess overfitting. This method, which examines the relationship of model performance over various numbers of training sample sizes, can help us to determine if a model is overfit and if it can benefit from more training examples [71].

We found a negative relationship between sample size and K-Fold-CV accuracies. Because increasing the number of training samples typically improves performance [72], this suggests that the lower estimates of accuracy from the larger samples are more likely to be correct than the higher estimates from smaller samples. Those higher estimates were likely biased as described in the prior section. Pulini et al [67] also reported a negative relationship of sample size and accuracies in ADHD imaging studies and Vabalas observed the negative relationship of sample size on reported accuracies in machine learning classifiers of autism spectrum disorders [73]. Both reviews were based on studies with sample sizes up to only ∼1,000. Similar observations were also made by Wolfers and colleagues when reviewing neuroimaging-based diagnostics for a number of different psychiatric disorders [70].

### Sample Heterogeneity and Data Imbalance

Although collaborative consortia, such as the ADHD-200, used relatively large samples sizes, such collaborations raise issues about sample heterogeneity and the use of imbalanced data. For example, like many other clinically referred samples, the ADHD-200 dataset had more boys than girls in the ADHD group compared with the control group. The ADHD group also had lower IQs than the control group. In addition, the demographic composition and sample acquisition methods differed across different study sites. The problem of dataset imbalance was addressed by several participating teams. Brown and colleagues from the University of Alberta found that models using only demographic information including age, sex, handedness, and IQ had sufficient statistical power to achieve a test accuracy 62.5%, higher than their models using fMRI features [57]. In the work of Colby et al. [74], a model using only demographic information had a higher accuracy (62.7%) than models using multimodal MRI features (55%). Both models using only the demographic features, although not meeting the requirements of the competition, outperformed the winning team that reported 61% accuracy using both structural and rs-fMRI data along with the demographic predictors in an ensemble model [43]. An additional study by Sidhu and colleagues also reported better accuracy using demographic information than the rs-fMRI features using the ADHD-200 dataset [75]. These observations highlight the concerns of data imbalance, and suggest that, if not dealt with carefully, the classifiers could be learning the neural correlates of the demographic features, rather than the diagnostic groups.

Some studies used methods to address the problem of unbalanced data. One approach is random undersampling, i.e, removing some research participants and creating a smaller sample size that is balanced for confounding factors [44, 45]. This is in contrast to oversampling, where some random samples from the minority classes were duplicated to create a lager and balanced dataset. Others used regression to control confounding factors such as age, sex, and acquisition sites, and used adjusted MRI features (residuals) in the classification algorithms [46, 47]. Some studies mentioned data balancing, but did not provide details on how it was done [48]. Lim et al [66] used a gaussian process classifier to discriminate 29 boys with ADHD from 19 control boys. The limited samples sizes prohibited subsampling to balance the data. They noted, although the boys with ADHD had significantly lower IQ than the control boys, the model-generated probability of having ADHD was not correlated with IQ, age, and other clinical features [66]. In more recent studies, more sophisticated methods such as Synthetic Minority Over-sampling Technique (SMOTE, [76]) were used to generate synthetic minority samples to combat the sample imbalance problem [49].

Previous studies from other fields have shown that not all the class balancing methods work equally well in reducing classifiers’ bias towards the majority class and guarantee good performance [77, 78]. In the ADHD studies that we reviewed, we did not find found higher held-out test accuracies in balanced studies than the unbalanced studies. Balanced studies in the K-Fold-CV group reported higher accuracies than those with unbalanced samples. However, they were mostly smaller studies than the unbalanced studies. It is not clear, at least for the K-Fold-CV group, if balanced designs led to higher accuracies, because sample size was a strong and negative predictor for the accuracies. Moreover, higher accuracies in the balanced LOOCV group were dependent on sample size (only significant for large studies with over 300 participants). This suggests that sample balancing may help performance in cases of large heterogenous studies. As expected, the results indicate that small sample sizes are not compensated by data balancing. More studies and larger sample sizes will be needed to find the appropriate class balancing methods and assess the potential benefit.

### Classification performance metrics

When test sets (or cross-validation sets) are also imbalanced, the overall accuracy may not be an ideal indicator of the performance of the classifier. A high accuracy can simply result from a classifier that classifies all samples into the class that has more participants. Most studies (N= 36, 75%) addressed this concern by also reporting sensitivity (True Positive rate, TP, the percentage of correctly identified cases (ADHD)) and specificity (True Negative rate, TN, the percentage of correctly identified controls). Three studies reported balanced accuracy, which is the arithmetic mean of the sensitivity and specificity; and three studies reported Youden’ s J-statistic (sensitivity + specificity -1).

Compared with percent correct, a better method to evaluate the overall performance of a classifier is the area under the Receiver Operating Characteristic curve (ROC [79]). The ROC curve plots sensitivity over the full range of false positive rates (equivalent to 1-specificity). The area under the ROC (AUC) measures the overall diagnostic accuracy of a classifier. Higher AUCs indicate better discriminating power (with 1 for the perfect classifier and 0.5 for the random non-discriminative classifier). The AUC is in general less sensitive to imbalance of a dataset compared with the percent correct measure, because AUC does not have bias toward models that perform well on the majority class at the expense of the minority class [78]. Davis et al [80] suggested that the area under the Precision-recall curve (AUPRC) is superior for assessing extremely imbalanced datasets and more informative than the ROC curve. The AUPRC plots precision (the percentage of examples classified as positive that are true positive, also known as positive predictive value, PPV) over recall (sensitivity). Overall, in the body of literature that we examined, no studies reported the AUPRC, and only 13 reported the AUC.

Other popular metrics for machine learning models are F1-score and Matthew’ s correlation coefficient (MCC). The F1-score is the harmonic mean of precision and recall. MCC is the correlation coefficient between the predicted and actual classes. Like the areas under the PRC or ROC, both the F1-score and MCC are better indicators of model performance than the percent correct statistic if test data classes are imbalanced. However, of the studies included in this review, only three reported F1-scores, and only two reported MCC.

Because most studies used percent correct to measures accuracy, we could only analyze percent correct in the current review. This may not represent the true model performance due to the limitations of this metric. We recommend that future studies adopt ROC or PRC analysis methods. Furthermore, inspecting the curves visually can reveal more information about how well the model discriminate classes at different decision thresholds. We don’ t recommend metrics such as the F1-score, MCC, and J-statistics, because these scores only capture the diagnostic matrix at a single threshold level. Furthermore, the performance metrics are important, not only for properly interpreting test results, but also for model training. If a model was trained by maximizing a biased metric, it will not be fully optimized to generalize to other samples. Finally, metrics that are insensitive to *class* imbalance (such as AUC or AUPRC) do not protect against biased due to feature imbalance as discussed in the prior section.

### Age and Sex

Although ADHD onsets prior to age 12, two-thirds of children continue to have symptoms and functional impairments into adulthood [81]. Longitudinal data show that some ADHD-associated brain alterations diminish during adolescence and adulthood [27, 29, 31]. Consistent with this, the very large ENIGMA ADHD study reported significant ADHD vs. control differences for children but not for adolescents and adults [34, 35]. Neuroimaging classifiers studies have focused on younger populations; only three studies developed ML classifiers for adults. Our observation of a lack of difference in predictive accuracy between classifiers for children/adolescents vs. adults is, therefore, inconclusive due to the small numbers of the adult studies. Few longitudinal studies have been reported for imaging in ADHD [27, 29, 31]. No machine learning models have been applied to longitudinal data yet. More efforts are needed to overcome the shortage of adult ADHD samples, as well as imaging data across the life span.

ADHD is more prevalent in boys than in girls [82, 83]. As a result, although the majority of the studies included samples from the both males and females, a high percentage of ADHD samples were from males (i.e., ∼80% male in ADHD-200 dataset). However, the control samples were often balanced (i.e., 52% male in ADHD-200 dataset). If sex is left unbalanced, it could result in erroneous prediction results, as we described in the above sections. Furthermore, brain alterations have been found to differ between the sexes at different ages [35, 84, 85]. The low representation of females in available samples may prevent the classifiers from learning female-specific brain alternations. Our quantitative analysis showed significantly higher accuracies in four male-only studies than other studies of sex-mixed samples. However, all four were studies with small sample sizes (<189), with three reporting LOOCV accuracies and the other reporting 10-Fold CV. Given the sample size effect and inflation by CV methods, it is inconclusive if ML models predict ADHD better in boys than girls.

### MRI Modality

Although we found no significant difference in the accuracies reported for the sMRI and rs-fMRI studies, the small number of studies using sMRI data preclude any meaningful inferences regarding which MRI modality is the most informative for discriminating ADHD patients from controls. Some studies attempted to identify the most informative MRI data modality. Qureshi et al [86] found that sMRI features yielded the highest prediction accuracy. Colby et al. [74] found that combined multi-modality features performed best compared with individual data modalities. However, all the MRI models performed worse than a classifier using only demographic features [74]. In a later study using a three-dimensional CNN model, Zou and colleagues extracted higher-level features from the sMRI and rs-fMRI modalities separately. This design leveraged the relationship between the two MRI modalities, yet still was able to extract independent features that collectively were useful for classification [87]. The authors also found that using multi-modal features outperformed either data modality alone [87]. Despite these individual observations, the overall lack of statistically significant differences in accuracies across different modalities in our review suggests that more studies are needed before we can determine which MRI modalities or combinations thereof are most informative for diagnostic classification.

### ML Classifiers

SVM was the ML model that was used most frequently, accounting for 38% of studies. SVM, however, is limited in handling images and relies on other preprocessing methods to extract a tabular representation of three-dimensional brain images. In more recent years, an increasing number of studies have used neurol networks [50, 51, 64, 88-90], particularly CNNs, which were developed for image analysis. We did not, however, observe statistically significant differences between the accuracies of the SVM and CNN models for ADHD. This finding is limited by the small number of studies using CNN classifiers. Nevertheless, because the use of CNNs will likely increase in the future, we here describe their current contributions to the field and their potential for the future.

Riaz et al. [91] used a CNN-based method (FCNet) to extract the functional connectivity (FC) of brain regions and then trained an SVM classifier using the extracted features to discriminate ADHD from control participants. The classifier achieved a highest held-out test accuracy of 68.6% for the ADHD-200 Peking subset. In the follow up study, the team built an end-to-end model system, DeepFMRI, which utilized multiple FCNets to extract features that were then fed into a deep neural network [92]. DeepFMRI streamlined the feature generation and selection as well as classification in one framework, and achieved a highest test accuracy of 73.1% for the NYU subset. Using preprocessed rs-fMRI and sMRI features as independent inputs, Zou et al. used a two-branched three-dimensional CNN to learn hierarchical features from each unique modality in a joint learning task. The multi-modal joint learning CNN architecture was superior to CNNs using either data modality alone [87]. Aradhya et al. [93] also used a CNN classifier and extracted features using the Deep Transformation Method (DTM).

Most studies, including many CNN studies, used pre-processed MRI features, such as those anatomized to an AAL template. Mao and colleagues argued that rather than using hand-crafted features, one should use a CNN to directly learn discriminatory features from images. Their four-dimensional CNN classifier, designed to learn and extract spatial and temporal features from rs-fMRI images, discriminated ADHD from control participants with an accuracy of 71.3% [94]. To increase their sample size and reduce overfitting, the authors augmented data by transforming rs-fMRI data into many short and fixed-lengthed video clips. Despite their promising results, they acknowledged that much work is still needed to localize the most discriminative sequences. Interestingly, a CNN using activation correlations from individual brain regions of the Default Mode Network (DMN) of the brain outperformed those using whole brain features [95]. Using only one relevant brain region substantially reduced feature space and complexity. The significantly improved model performance also suggests that current sample sizes, in relation to the number of features available, maybe limiting the CNN models’ capacity.

With more samples becoming available in the future, and the increased datasets of publicly available raw MRI images, CNN methods will likely to be seen in more and more studies and be explored to their full capacity for feature extraction and classification as has been the case for computer vision [96-100].

### Building Larger Datasets

Sample size has been a major bottleneck impeding the development of more accurate and clinically useful imaging classifiers for ADHD. The largest MRI dataset, to date, has been built by the Enhancing Neuro Imaging Genetics Through Meta-Analysis (ENIGMA) consortium. Under the umbrella of the ENIGMA consortium, many independent working groups for specific diseases or phenotypes have been established, including ADHD. By implementing standardized data processing protocols and pipelines, the ENIGMA consortium made it possible to share data across many sites to perform within-disorder and cross-disorder studies [101-104]. The ENIGMA ADHD Working Group has obtained over 4,100 samples of ADHD participants and controls from 37 sites thus far. In the initial ENIGMA ADHD reports, Hoogman and colleagues reported that, for children, ADHD was associated with significant volumetric reductions in intracranial volume, amygdala, caudate nucleus, nucleus accumbens, hippocampus, and cortical surface areas from many regions [34, 35]. No significant differences were found for adolescents or adults. Furthermore, the estimated effect sizes for children were small, ranging from 0.11 to 0.19. Users of the ENIGMA ADHD dataset, however, face the same problems of data heterogeneity and imbalanced demographic groups as those using the ADHD-200 dataset. Significant challenges remain when using such data to build a machine learning classifier. Furthermore, the ENIGMA ADHD data is primarily preprocessed sMRI data in tabular form. Not all sites have data on other modalities, such as rs-fMRI or DTI, available for their samples. The ENIGMA ADHD sites have not yet pooled raw MRI images, which is needed for CNN models. Nevertheless, we encourage the research community to continue to contribute ADHD samples to this cohort and provide more open access to various data modalities. Future studies that attempt to advance ML-powered ADHD diagnostic classifiers should use the ENIGMA ADHD dataset available, as this resource has been under-utilized in AI and ML studies of ADHD.

## Conclusions

Our review of ML studies of MRI-based ADHD diagnostic classifiers has important implications for methods development, but these studies have not yet led to clinically useful classifiers. Our review shows that the variability of results across studies is due, in part, to differences in methodology. Future work should use the largest samples possible and should rely on a held-out test set, rather than cross-validation for estimating prediction accuracy. Future studies should not rely on percent correct as a measure of accuracy in unbalanced samples. Our analysis also highlighted the need of data from underrepresented groups, particularly females and adults. We hope that our review provides a better understanding of the efforts invested in developing ADHD imaging classifiers in the field and encourages more stringent model design and data processing for future studies. In the meanwhile, the initial results from the ENIGMA ADHD consortium should encourage more sites to participate. The lack of a very large multi-modal dataset that include sufficient data from both sex and all ages may be the biggest impediment to developing a clinically useful classifier for diagnosing ADHD.

## Data Availability

NA

## Acknowledgement

Dr. Zhang-James is supported by the European Union’ s Seventh Framework Programme for research, technological development and demonstration (grant number 602805) and the European Union’ s Horizon 2020 research and innovation programme (grant number 965381).

Dr. Hoogman is supported by a personal Veni grant from of the Netherlands Organization for Scientific Research (NWO, grant number 91619115)

Dr. Franke is supported by a personal Vici grant (grant number 016-130-669) from the Netherlands Organization for Scientific Research (NWO).

Dr. Faraone is supported by the European Union’ s Seventh Framework Programme for research, technological development and demonstration (grant number 602805), the European Union’ s Horizon 2020 research and innovation programme (grant number 965381 & 728018) and United States National Institute of Mental Health (NIMH, grant numbers 5R01MH101519 and U01 MH109536-01).

## Supplementary Figures

**Supplementary Figure 1.**
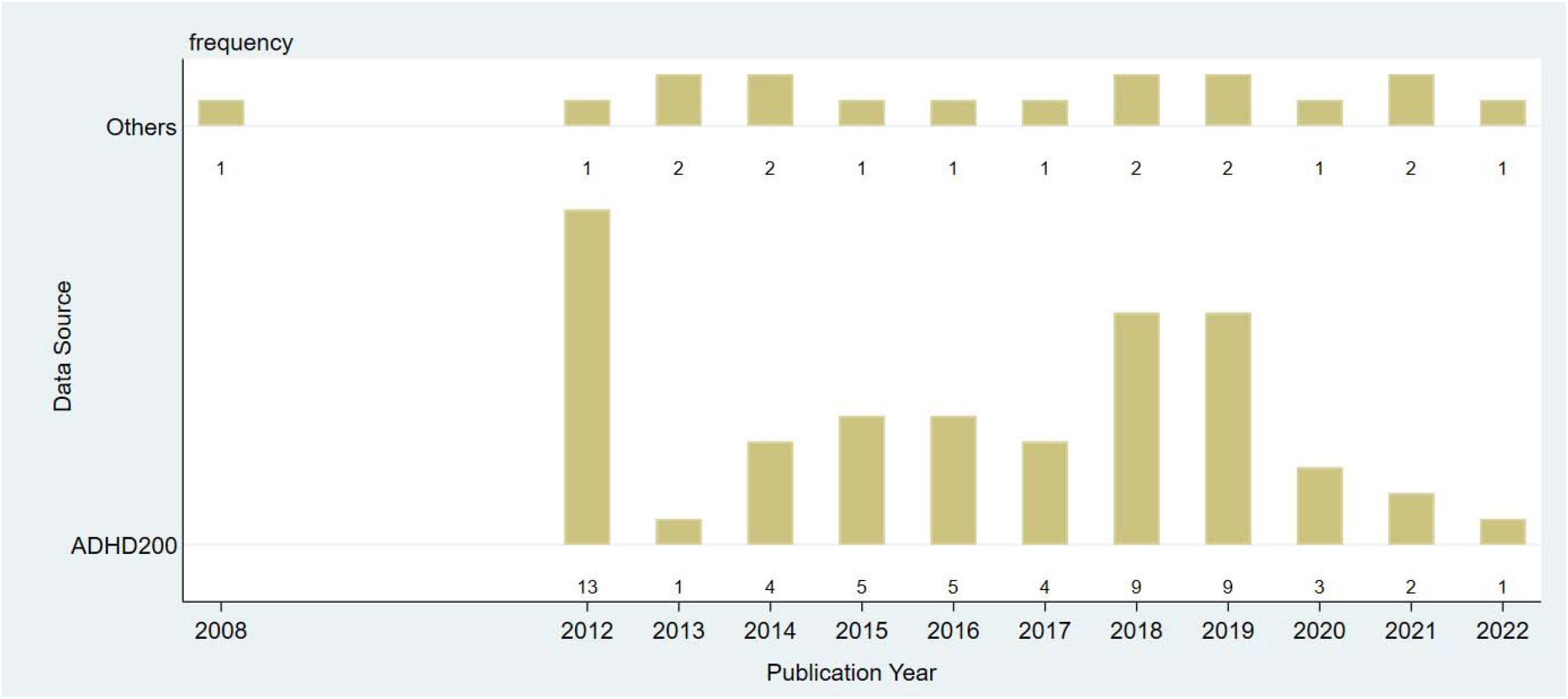
Numbers of publication in each year. The top row includes studies that used non-ADHD-200 data; the bottom row includes studies that used ADHD-200 data. The numbers for each year are labeled beneath the bar.

**Supplementary Figure 2.**
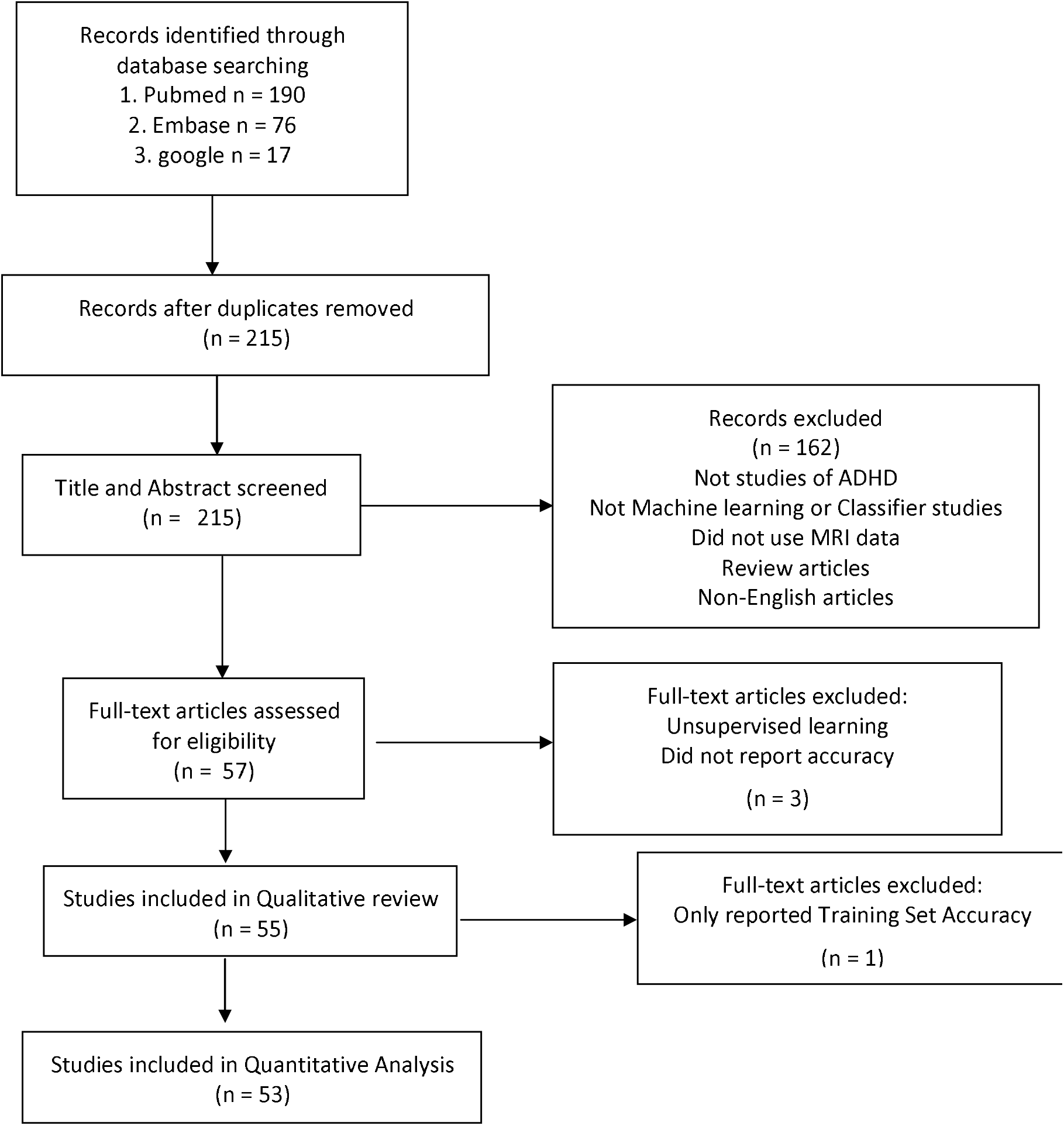
PRISMA flow diagram for review and meta-analysis.

**Supplementary Figure 3.**
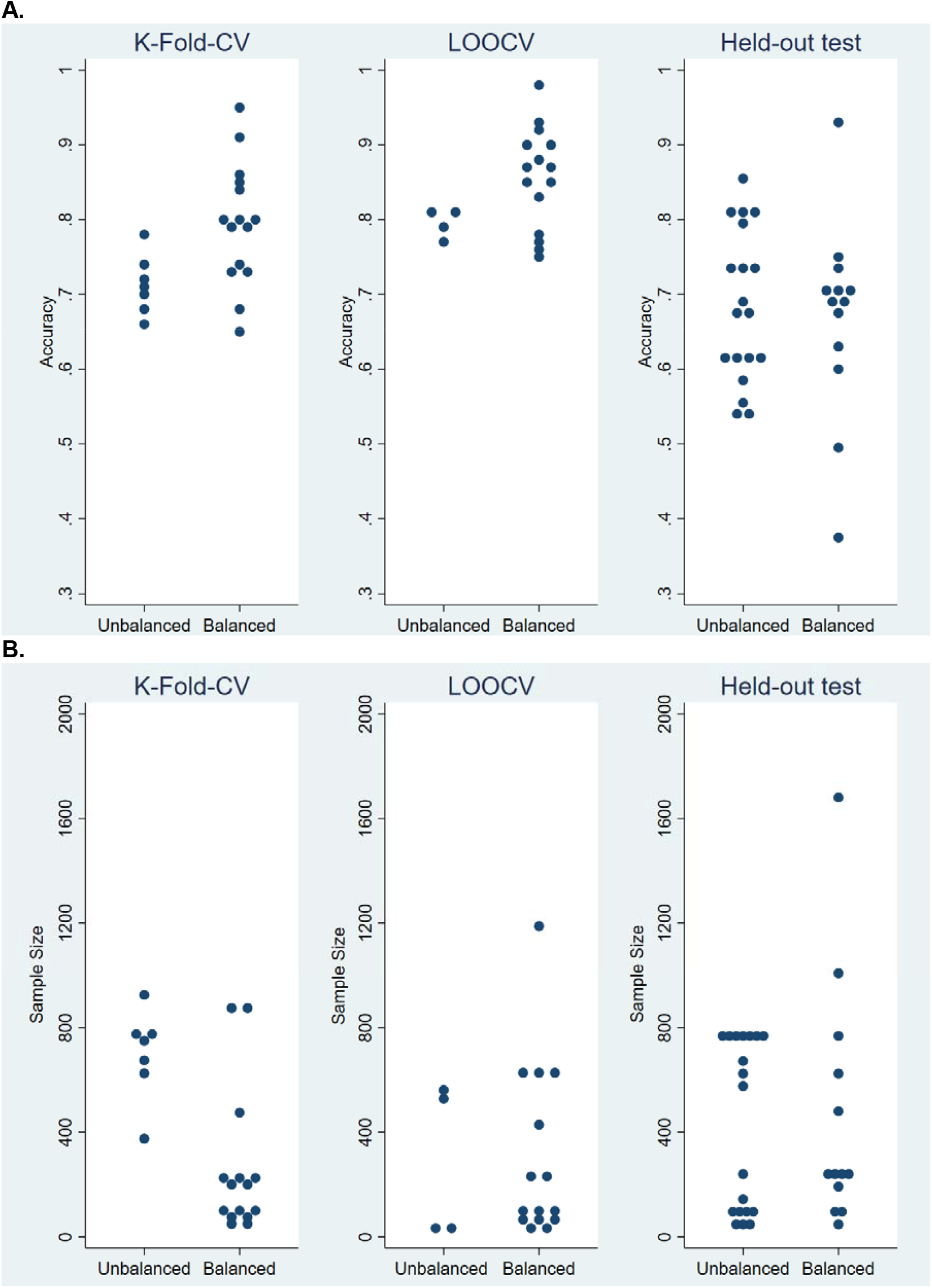
A. Reported accuracies and training data balancing. B. Training sample sizes in balanced vs unbalanced studies.

**Supplementary Figure 4.**
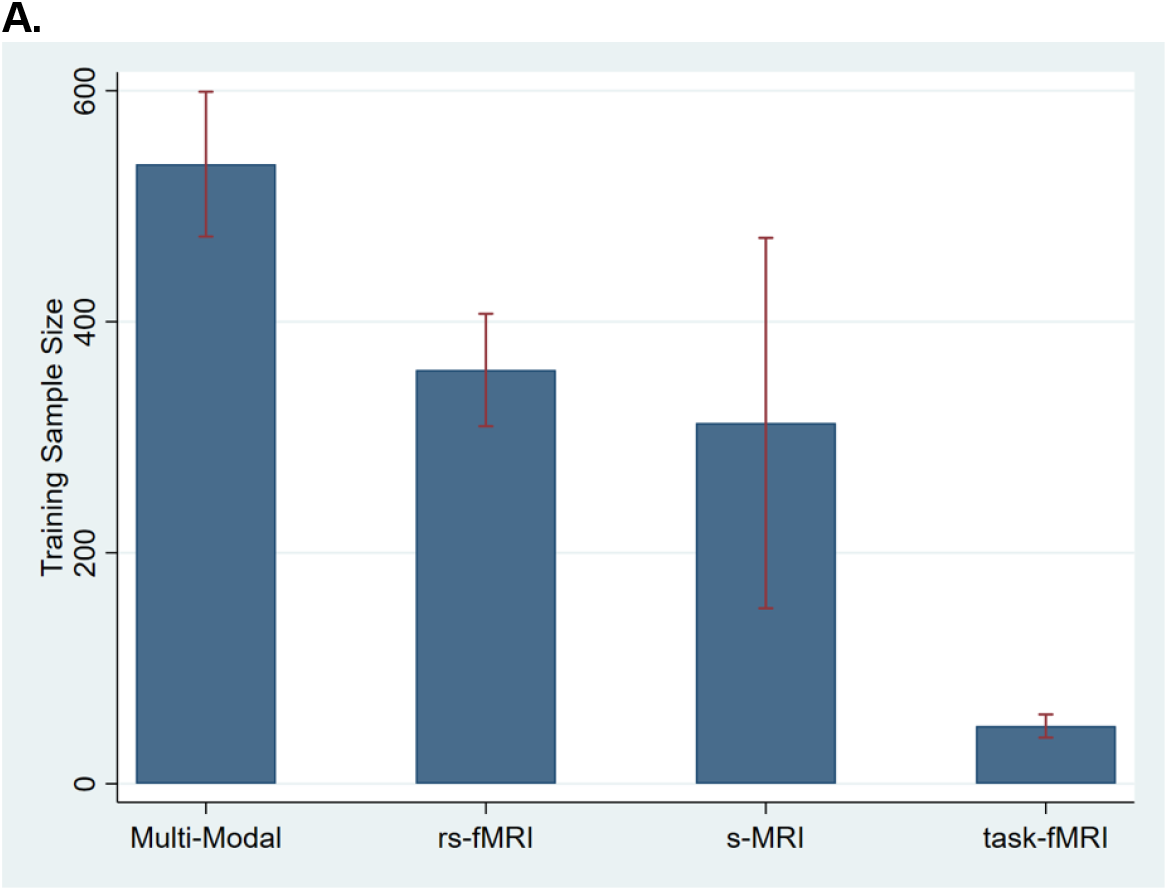
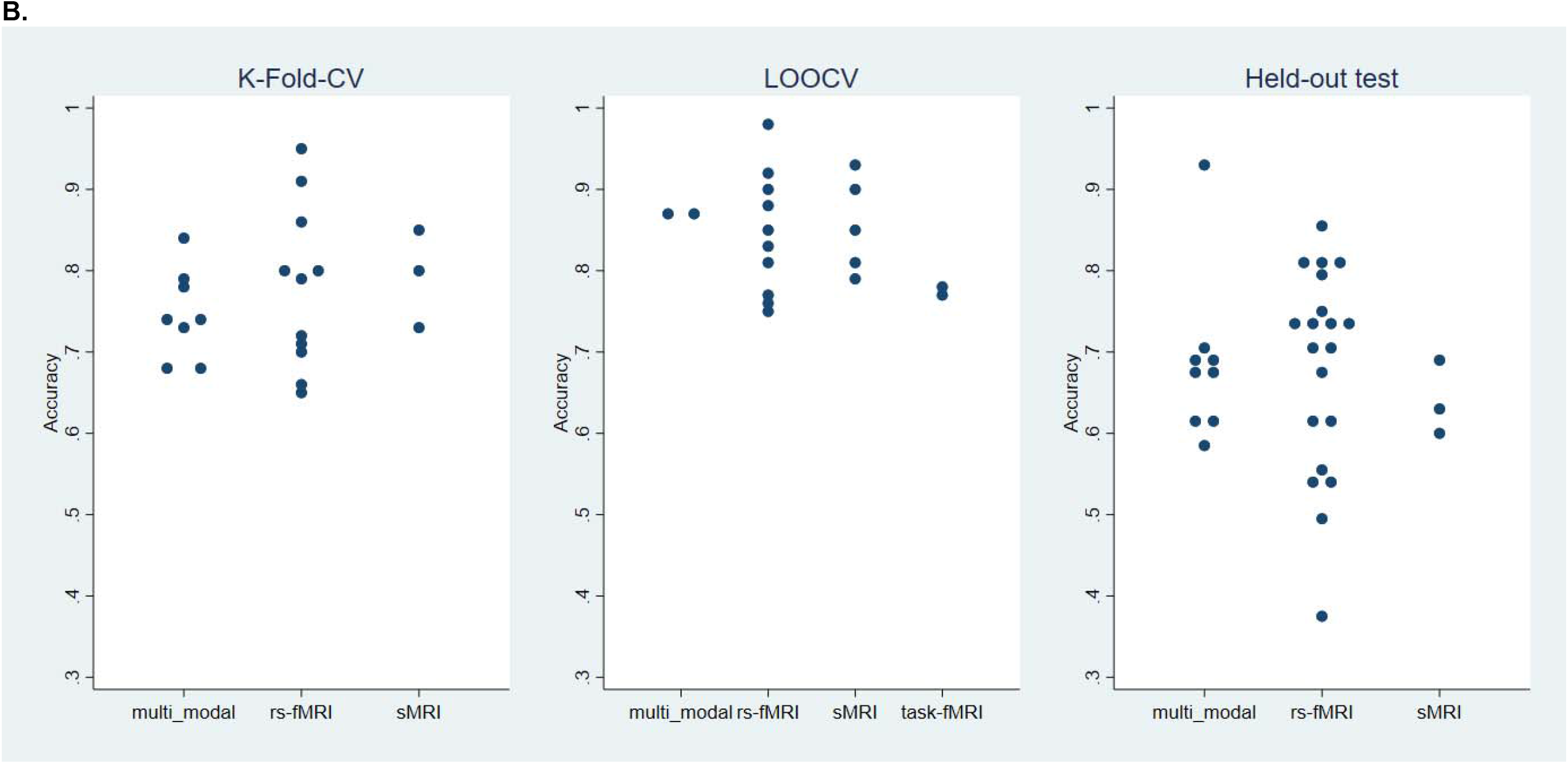
A. Mean and standard errors of the sample size for multi-modality, rs-fMRI, sMRI and task-based fMRI studies. B. Accuracies in studies using different MRI modalities.

**Supplementary Figure 5.**
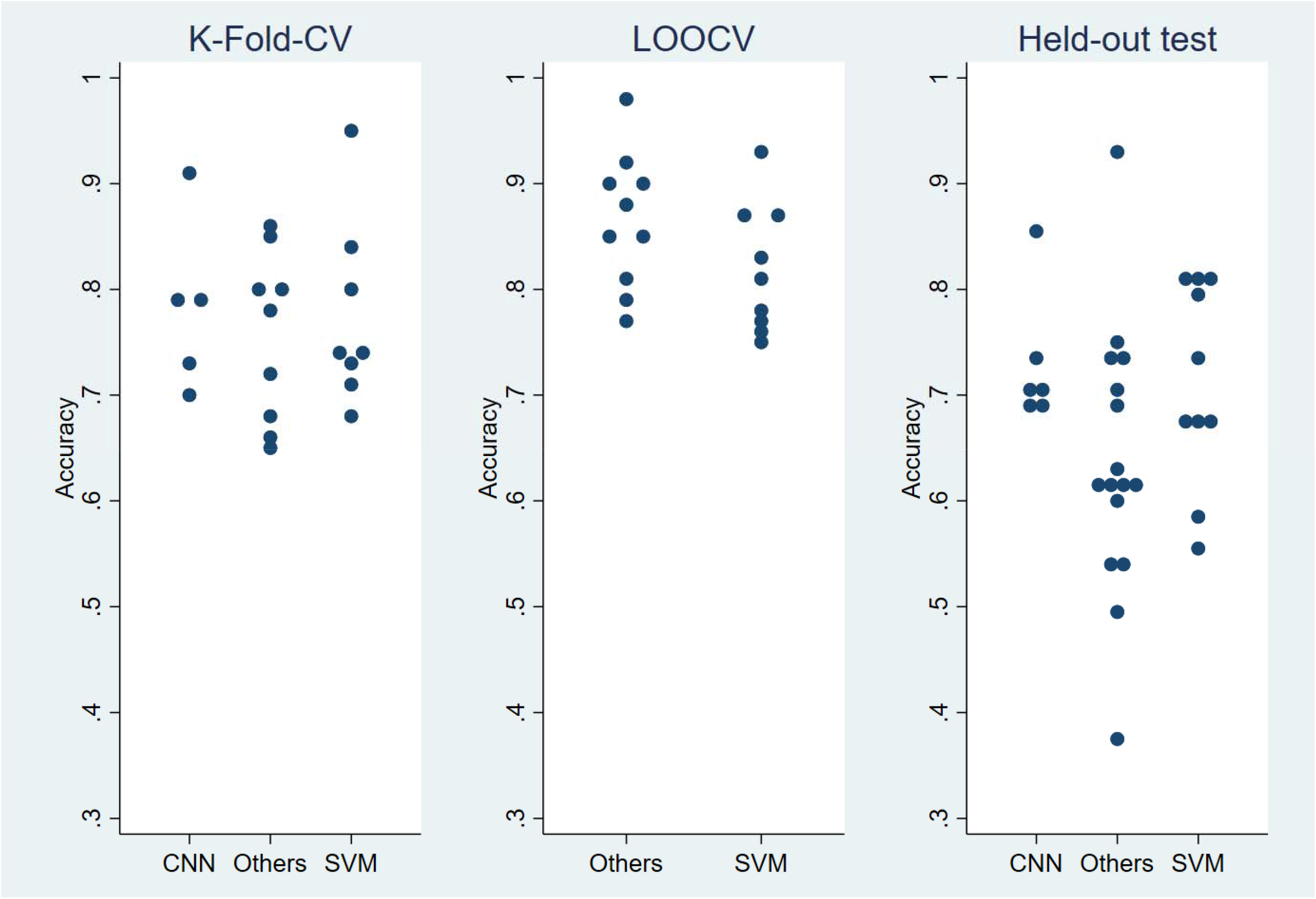
Accuracies in studies using different ML models.

